# Angiotensin-converting enzyme inhibitors and angiotensin receptor blockers are not independently associated with lower pericoronary adipose tissue attenuation on CT in patients with type 2 diabetes mellitus

**DOI:** 10.64898/2025.12.04.25341670

**Authors:** Bryan Wu, Hanyi Joh, Koen Nieman, Ryan Sandoval

**Author notes:** Corresponding author: Bryan Wu, M.D., 300 Pasteur Drive, Palo Alto, CA 94305.

## Abstract

**Background:** In patients with type 2 diabetes mellitus (T2DM), angiotensin-converting enzyme inhibitors (ACE-I) and angiotensin receptor blockers (ARB) are first-line antihypertensive treatments with important cardiovascular benefits and their impact on coronary-specific inflammation is unknown. Pericoronary adipose tissue (PCAT) attenuation, as assessed by coronary computed tomography angiography (CCTA), serves as specific biomarker for coronary inflammation and we aim to assess whether treatment with ACE-I or ARB is correlated with lower PCAT attenuation.

**Methods:** In this retrospective observational study, we analyzed 223 patients with T2DM and coronary atherosclerosis who underwent CCTA between 1/1/2017 through 9/1/2024 at our institution. PCAT attenuation was measured in the proximal right coronary artery. Multivariate linear and logistics regression analyses were performed.

**Results:** Of the 223 patients (mean age 64.9±8.8 years, 69.1% male), 122 patients were on ACE-I or ARB (ACE-I/ARB). ACE-I/ARB users were more likely to be on high-intensity statin therapy, which is known to stabilize plaques and reduce coronary inflammation (44.3% vs 23.8%, p=0.001). Treatment with ACE-I/ARB was not independently associated with lower PCAT attenuation (-73.5±7.3 HU vs. -71.6±8.0 HU; adjusted p=0.478), Subgroup analysis revealed lower PCAT attenuation in ACE-I/ARB users with glomerular filtration rate (GFR) <90 mL/min (-74.8±6.6 vs. -71.4±7.1 HU; p=0.038). Other antihypertensives (beta blockers, calcium channel blockers, thiazides) were not linked with lower likelihood of elevated coronary inflammation (PCAT attenuation ≥-70.5 HU).

**Conclusions:** In T2DM patients with coronary atherosclerosis, ACE-I/ARB use was not independently associated with lower coronary inflammation as defined by PCAT attenuation, though benefits may exist in those with reduced GFR.

**Clinical Perspective:** *What is New?:*

- In patients with type 2 diabetes mellitus and coronary atherosclerosis, treatment with angiotensin-converting enzyme inhibitors and angiotensin receptor blockers is not independently associated lower pericoronary adipose tissue attenuation, a marker of coronary inflammation on CT, but such benefit may exist in patients with glomerular filtration rate <90 mL/min.
- Other classes of antihypertensive medications (beta blockers, dihydropyridine calcium channel blockers, and thiazides) were also not associated with lower coronary inflammation.

*What are the Clinical Implications?:*

- Unlike the high-intensity statins, no particular class of antihypertensive medications was shown to possess anti-inflammatory potential in coronary plaque stabilization but further studies are warranted to see whether patients with renal dysfunction would benefit more from angiotensin-converting enzyme inhibitors and angiotensin receptor blockers in this clinical context.

## Introduction

Angiotensin receptor blockers (ARB) and angiotensin-converting enzyme inhibitors (ACE-I) are first-line antihypertensive therapies for patients with type 2 diabetes (T2DM), and there have been several studies demonstrating the cardiovascular benefits linked with renin-angiotensin-aldosterone system (RAAS) inhibitors in this high-risk cohort.^1;2^ Earlier studies have shown that angiotensin II can lead to a higher production of proinflammatory cytokines, reactive oxygen species, and adhesion molecules.^3;4^ Its involvement in coronary atherogenesis has been suggested as patients with critical coronary artery disease have higher plasma levels of angiotensin II.^5^ While ACE-I and ARB have demonstrated anti-inflammatory potential by reduction of systemic biomarkers, such as C-reactive protein and interleukin-6,^6^ it is unknown whether ACE-I or ARB can modulate inflammation specific to the coronary artery system.

Pericoronary adipose tissue (PCAT) attenuation is a novel imaging biomarker for coronary inflammation, which is associated with plaque progression and rupture. As assessed by coronary computed tomography angiography (CCTA), PCAT is the epicardial adipose tissue that is directly adjacent to the coronary vessel. Heightened inflammation from the coronary plaque impairs the maturation of the adipocytes and reduces their sizes.^7^ This intricate process ultimately leads to elevated density of adipocytes of the PCAT, which corresponds to higher Hounsfield Units (HU) on CCTA. A mean PCAT attenuation above -70.5 HU in the proximal right coronary artery (RCA), which has been the traditional site of assessment, is predictive of future myocardial infarction with a hazard ratio of 2.45, and this biomarker has been shown to outperform CT calcium score, cardiovascular risk score, and even the presence of obstructive lesions in predicting future myocardial infarction.^8^

To our knowledge, there has been no study that closely assessed the effect of antihypertensive therapies on PCAT attenuation, especially in patients with T2DM. T2DM and vascular inflammation are closely linked where hyperglycemia promotes increased production reactive oxygen species and secretion of pro-inflammatory cytokines.^9^ In our prior study, we found that treatment with high-intensity statin therapy was independently associated with lower PCAT attenuation in T2DM patients with coronary atherosclerosis.^10^ In this retrospective analysis where we expanded our sample size from the aforementioned study, we aim to evaluate whether treatment with ACE-I or ARB is correlated with lower PCAT attenuation in this high-risk cohort.

## Methods

### Patient screening and clinical data collection

In this retrospective observational study, we screened patients who underwent CCTA from January 1, 2017 through September 1, 2024 at Stanford Health Care. To qualify for the diagnosis of T2DM and hypertension, patients must have the diagnosis clearly documented in a physician note. Additionally, for T2DM, patients need to have prior fasting glucose ≥126 mg/dL, hemoglobin A1C (HbA1C) ≥6.5%, random glucose ≥200 mg/dL, or a blood glucose level of ≥200 mg/dL on a 2-hour oral glucose tolerance test. For hypertension, patients must either receive antihypertensive treatment for ≥6 months or have a documented blood pressure ≥130/80 mmHg during 2 or more clinic visits. The inclusion criteria are: age 40-80 years, diagnosis of T2DM with most recent hemoglobin A1C (HbA1C) ≤ 9.0%, body mass index (BMI) ≤ 35.0, and evidence of coronary atherosclerosis on CCTA (as defined by Coronary Artery Disease-Reporting and Data System [CAD-RADS] grading ≥1), computed tomography (CT) tube voltage 100–120 kV, and treatment with at least 1 oral antidiabetic agent for ≥6 months. We excluded patients who had suboptimal imaging quality, prior coronary interventions and anomalous coronary anatomy including non-dominant right coronary artery. The majority of our patients did not previously undergo coronary imaging. We also excluded patients with obstructive lesions in the right coronary artery as these findings could raise the PCAT attenuation in that region and thus misrepresent the global inflammation for the entire coronary system.

We collected patients’ demographic information, atherosclerotic risk factors, and medication history. Patients’ medication regimens were verified for analyses if the treatment duration was greater than 6 months. Additionally, we also recorded patients’ blood pressures at the time of CCTA acquisition (before beta blockers and nitroglycerin were administered) and during the 2 outpatient clinic encounters within 6 months prior to scan acquisition if available. Laboratory data, including lipid panel and basic metabolic panel, were also documented. The glomerular filtration rate was calculated the 2021 Chronic Kidney Disease Epidemiology Collaboration (CKD-EPI) equation using sex, age, and serum creatinine.

### CT imaging acquisition and PCAT analysis

The CCTAs were acquired on a third generation, dual-source, 384 (2 × 192) slice CT scanner (Siemens Somatom FORCE, Forchheim, Germany) with retrospective EKG-gating. Patients were instructed to perform a breath-hold during the imaging acquisition. Oral beta blockers were administered if their resting heart rate exceeded 70 beats per minute and sublingual nitroglycerin was given to all patients to allow for coronary dilatation. The severity of coronary artery disease as defined by the CAD-RADS classification system and coronary artery calcium (CAC) scores were recorded. The PCAT analysis was done on the Terarecon Aquarius Intuition 4.4.13 software by a cardiologist who is board-certified in cardiac CT and blinded to patients’ clinical profiles. For majority of the scans, end-diastolic phase images were used for analysis. The PCAT attenuation was measured in the proximal RCA, starting from 1cm distal to the coronary origin along a 4-cm length, and the width is defined as the diameter of the coronary vessel (Figure 1). While there is no universal definition for where PCAT attenuation should be measured, this site is commonly used due to its lack of side branches and abundance of adipose tissue.^1^ Within this region of interest, the PCAT attenuation is defined as mean HU attenuation within the range for adipose tissue on CT (-190 to -30 HU). As described in prior study, the PCAT attenuation was divided by a correction factor of 1.11485 for CCTAs acquired with tube voltage of 100 kV.^9^

**Figure 1.**
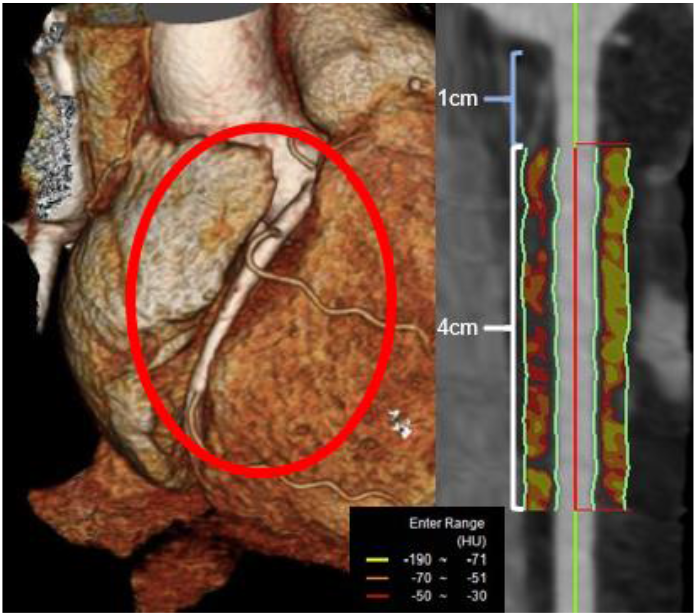
Illustration of a patient’s pericoronary adipose in the proximal right coronary artery (1cm away from the origin, along a 4cm length). Red indicates more inflamed adipose tissue.

### Statistical analysis

IBM SPSS Statistics 30.0.0 software was used for statistical analysis. Continuous variables are presented as mean ± standard deviation, while categorical variables are presented as percentages of patients. Univariate analyses were conducted with unpaired student’s t-test or Fisher’s exact test. Multivariate linear regression analysis (adjusting for age, sex, BMI, hyperlipidemia, history of tobacco use, microvascular complications, presence of obstructive coronary lesions requiring revascularization, HbA1C, treatment with high-intensity statin therapy and glucagon-like peptide-1 receptor agonists) was conducted to evaluate the difference in PCAT attenuation in patients who were on ACE-I or ARB against their counterpart. Multivariate logistics regression model, adjusting for the aforementioned variables, was used to determine whether the different classes of first-line antihypertensive therapies are associated with lower likelihood of elevated coronary inflammation (PCAT attenuation ≥ -70.5 HU). For our subgroup analyses, the cutoffs for HbA1C, GFR, and blood pressure were made based on clinical relevance and evenness in distribution of patients.

This study was reviewed and approved by the Stanford University Institutional Review Board and informed consent was waived.

## Results

As illustrated in Figure 2, a total of 1223 patients were screened, and 223 patients met our study criteria and were included in the study. 122 patients were on either ACE-I or ARB (ACE-I/ARB), with 55 patients on ACE-I and 67 patients on ARB. 101 patients were not on either antihypertensive therapy. The mean age for our study cohort was 64.9±8.8, and majority of our patients were male (69.1%). The ACE-I/ARB group had higher CAC score (759.5±1026.1 vs 518.8±781.3; p=0.048) and higher prevalence of hypertension (97.5% vs 75.2%; p<0.001) and hyperlipidemia (92.6% vs 82.2%; p=0.023). Chest pain evaluation was the common indication for both groups (ACE-I/ARB: 64.8%, not on ACE-I/ARB: 57.4%; p=0.272). ACE-I/ARB group had higher proportion of patients on high-intensity statin therapy, which is known to stabilize coronary plaques and reduce inflammation (44.3% vs 23.8%, p=0.001).

**Figure 2.**
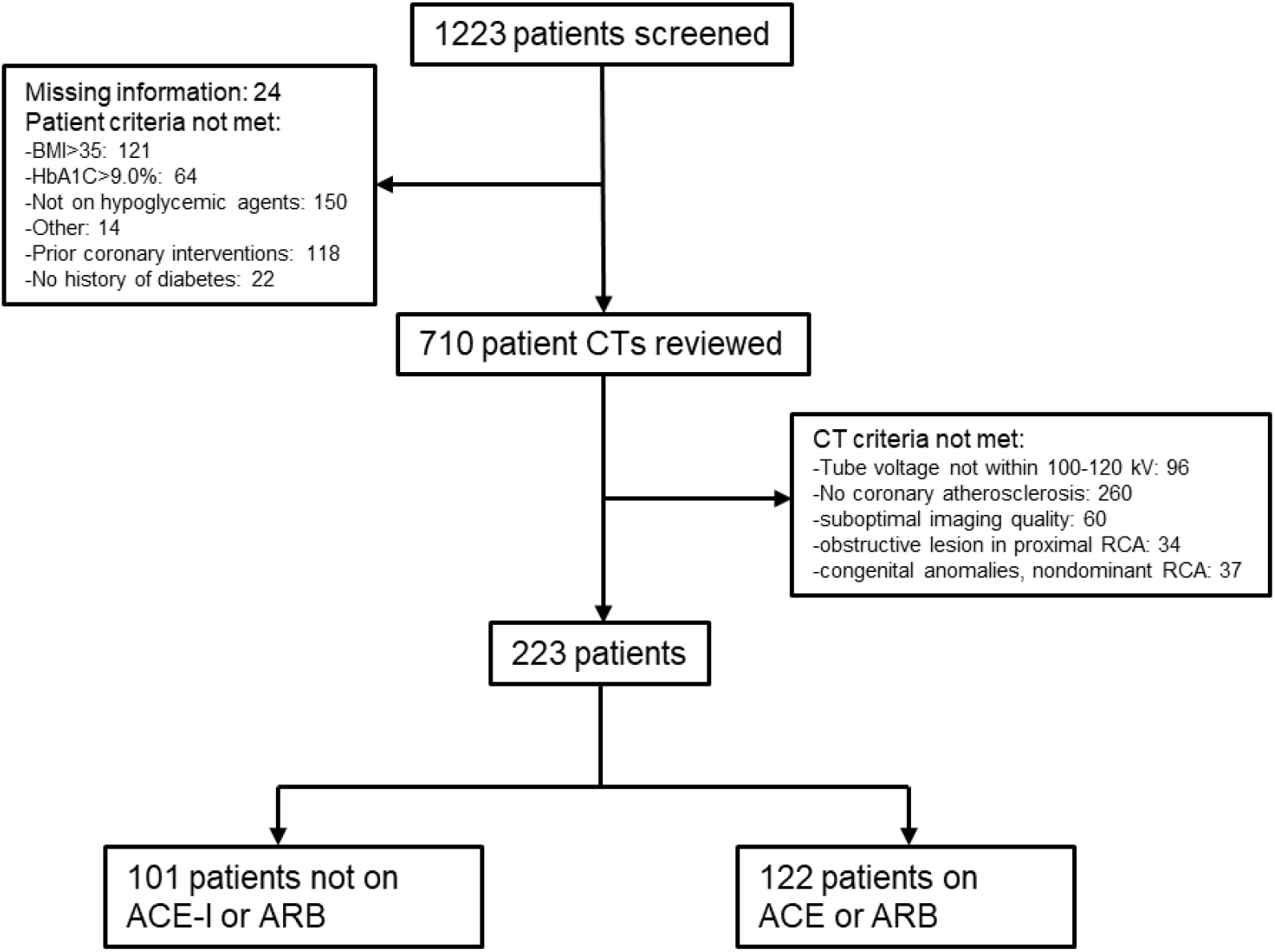
Diagram of patient screening and selection process. Acronyms: BMI: body mass index, HbA1C: hemoglobin A1C, ACE-I: angiotensin-converting enzyme inhibitors, ARB: angiotensin receptor blockers, RCA: right coronary artery, CT: computed tomography.

In the comparison of PCAT attenuation between the three groups, both univariate and multivariate linear regression analyses were performed. As shown in Figure 3, patients on ACE-I/ARB had non-significantly lower PCAT attenuation (-73.5±7.3 HU, -71.6±8.0 HU, unadjusted p= 0.073; adjusted p=0.478). Patients on ACE-I had similar PCAT attenuation as those on ARB (-73.3±7.0 HU vs -73.6±7.6 HU, respectively; unadjusted p=0.808, adjusted p=0.985).

**Figure 3.**
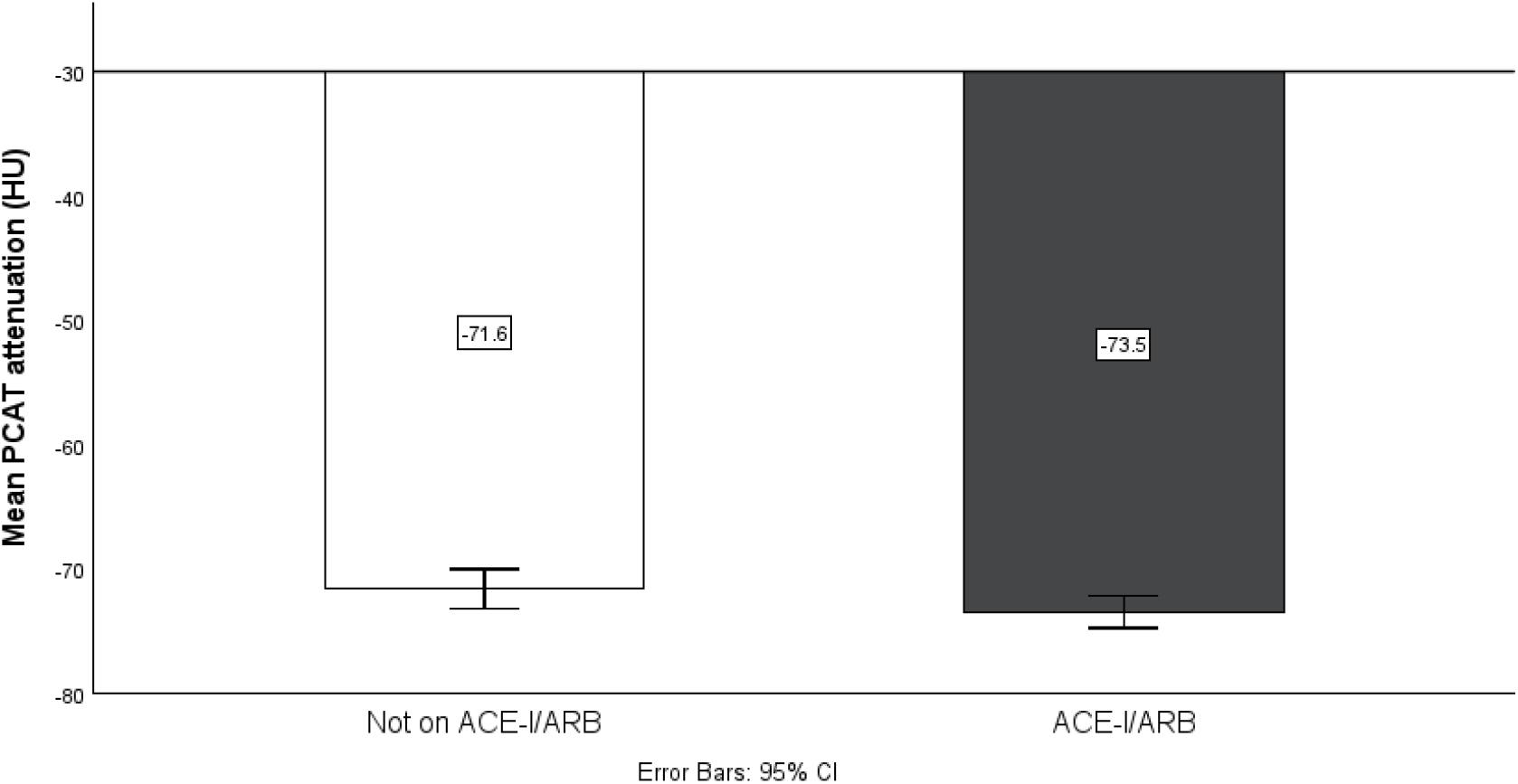
Comparison of pericoronary adipose tissue (PCAT) attenuation between patients on angiotensin-converting enzyme inhibitors (ACE-I) or angiotensin receptor blockers (ARB) and those not on either treatment.

The results for our subgroup analyses are illustrated in Figure 4, and comparisons were made with adjustment for high-intensity statin treatment. 97% of patients had their creatinine and GFR assessed in the 6 months prior to CCTA. In patients with GFR<90 mL/min, those who were treated with ACE-I or ARB had lower PCAT attenuation (-74.8±6.6 HU vs -71.4±7.1 HU, p=0.038). In this subgroup, we also assessed the effect of sodium-glucose cotransporter-2 (SGLT-2) inhibitors (n=28), another class of medications with renal protective properties, but found no difference (SGLT-2 inhibitors: -73.5±6.4 HU, not on SGLT-2 inhibitors: -73.4±7.2 HU; p=0.997). We had a small number of patients with GFR<60 mL/min (n=24) and noted a non-significant trend towards lower coronary inflammation associated with ACE-/ARB (-76.6±7.4 HU vs -70.9±6.7 HU; p=0.061). Finally, among patients already on high-intensity statins, ACE-I/ARB users did not have lower PCAT attenuation.

**Figure 4.**
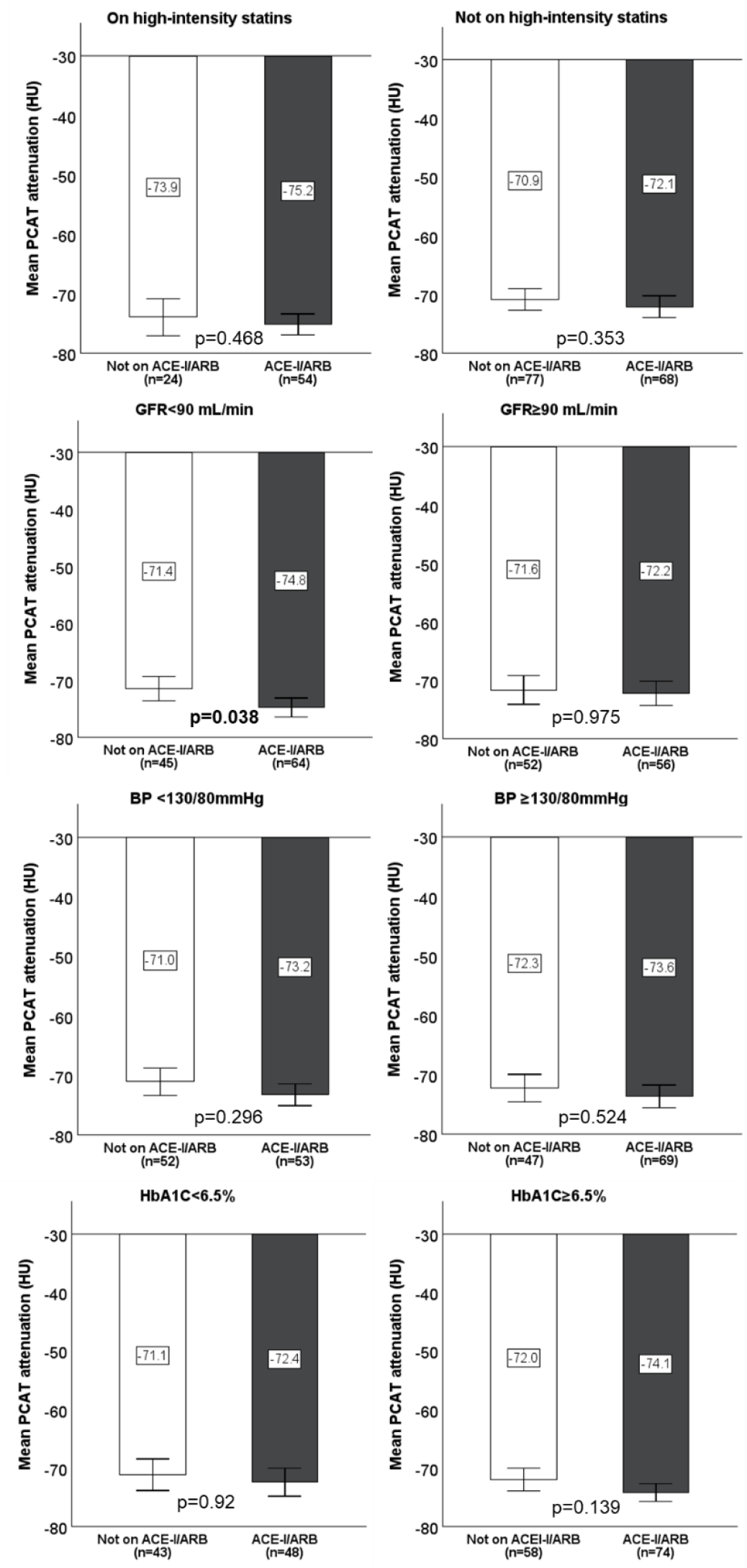
Subgroup analysis of PCAT attenuation in patients on either angiotensin-converting enzyme inhibitors (ACE-I) or angiotensin receptor blockers (ARB) versus their counterpart. Comparisons were made with adjustment to high-intensity statin treatment except for the first subgroup. Acronyms-HbA1C: hemoglobin A1C, CAD-RADS: Coronary Artery Disease Reporting and Data System.

We also performed a multivariate logistics regression for other classes of antihypertensive regimen to identify independent predictors for elevated PCAT attenuation (≥-70.5 HU). Overall, ARB was the most frequently prescribed antihypertensive medication (30%), followed by ACE-I (24.7%) and beta blockers (24.7%). We did not include aldosterone antagonists, loop diuretics, non-dihydropyridine calcium channel blockers, and other classes of antihypertensive therapies as there were fewer than 30 patients on each of those drug classes. As shown in Figure 5, none of the antihypertensive medications had lower likelihood of heightened coronary inflammation. We also found that patients with elevated PCAT attenuation were on similar number of antihypertensive therapies as their counterpart (1.2±1.1 vs 1.4±1.0, respective, p=0.156). Furthermore, these two groups had similar systolic blood pressure (elevated PCAT: 131±14 mmHg; non-elevated PCAT: 130±14 mmHg; p=0.875) and patients with elevated coronary inflammation had non-significantly higher diastolic blood pressure (76±28 mmHg vs 72±11 mmHg, p=0.118).

**Figure 5.**
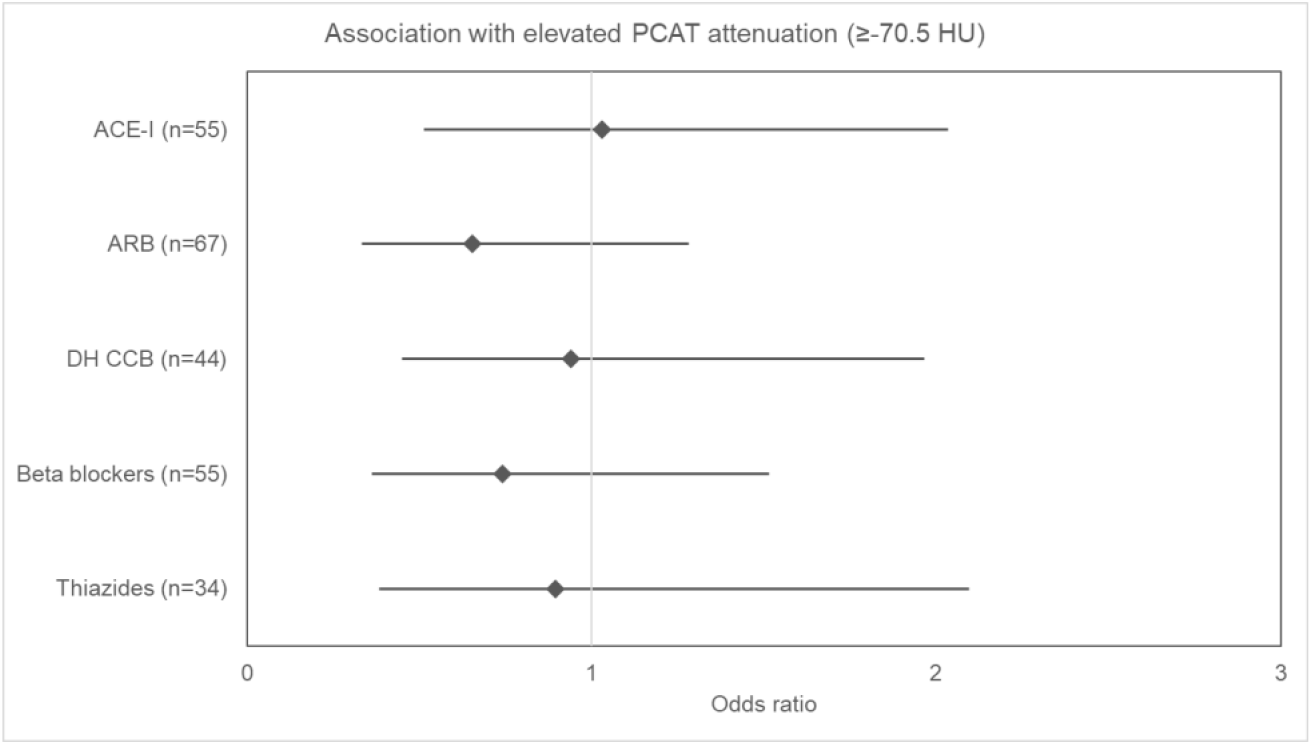
None of antihypertensive treatment classes were associated with lower likelihood of elevated coronary inflammation (pericoronary adipose tissue [PCAT] attenuation ≥-70.5 Hounsfield Unit). Each bar represents 95% confidence interval of odds ratio obtained by multivariate logistics regression model. Acronyms: ACE-I: angiotensin-converting enzyme inhibitor, ARB: angiotensin receptor blocker, DH CCB: dihydropyridine calcium channel blockers.

## Discussion

To our knowledge, this is the first study that evaluated the impact of various classes of antihypertensive therapy on coronary inflammation by PCAT attenuation in patients with type 2 diabetes mellitus who are high risk for vascular inflammation. We found that those who were treated with ACE-I or ARB for greater than 6 months were more likely to be on high-intensity statin therapy, which has shown to yield plaque stabilization and lower coronary inflammation,^10; 11^ and treatment with these classes of drugs was not independently associated with lower PCAT attenuation on CCTA.

While RAAS inhibitors are shown to lower serum biomarkers for inflammation,^6^ evidence supporting their prevention of future myocardial infarction through stabilization of coronary plaque progression and vulnerability is less robust. In a recent retrospective study by Zhang et al. of 2501 patients with hypertension and diabetes who were diagnosed with obstructive coronary artery disease on coronary angiogram, those who were on ACE-I/ARB before diagnosis had lower composite major adverse cardiac and cerebral events, but did not have lower incidences of non-fatal MI.^12^ In the Quinapril Ischemic Event trial, treatment with quinapril did not reduce ischemic events and also yielded no significant impact on angiographic progression of coronary plaques.^13^ On the contrary, in the OLIVUS trial, treatment with Olmesartan was shown to yield slower plaque progression, as assessed by intravascular ultrasound, in nonculprit vessels in patients undergoing percutaneous coronary intervention.^14^ Nevertheless, a more recent sub-analysis of the PARADIGM registry showed that ACE-I and ARB had no significant longitudinal effect on plaque volume and composition.^15^

In our subgroup analyses, we found that among patients with GFR lower than 90 mL/min, ACE-I/ARB users had lower PCAT attenuation. When renal perfusion and GFR are reduced, there is an amplified RAAS activity from increase in renin production from juxtaglomerular cells that line the afferent arterioles.^16^ As increase in production of angiotensin II has shown to induce vascular remodeling through promotion of endothelial dysfunction and oxidative stress,^17^ it is plausible that patients with lower GFR and more heightened RAAS activity at baseline would receive greater anti-inflammatory benefit from ARB and ACE-I. In the RENAAL study, there was a trend towards lower incidence of MI in patients with type 2 diabetes and nephropathy who were treated with losartan.^18^ In an observational study of patients who experienced an acute myocardial infarction in the SWEDEHEART registry, those with GFR between 30-60 mL/min had more pronounced benefit from RAAS inhibitors in prevention of recurrent MI than those with GFR above that cutoff. Our study had a limited number of patients with GFR<60 mL/min (n=24) as medical providers would likely have elected for alternative ischemic evaluation modalities that do not require intravenous contrast in patients with more advanced kidney dysfunction. We observed a trend towards lower PCAT attenuation in patients on RAAS inhibitors for this particular subgroup (p=0.061), but the findings need to be further assessed on a larger-scale population. We were also unable to fully account for the role of microalbuminuria as only 27% of patients had this data available, which is a limitation.

In patients with chronic coronary disease, the 2023 American Heart Association guidelines assigned a stronger class of recommendation for RAAS inhibitors in those with other comorbidities including hypertension, diabetes, heart failure with reduced ejection fraction (<40%) and chronic kidney disease compared to those who do not have the aforementioned conditions.^19^ As our findings are not suggestive of anti-inflammatory potential associated with RAAS inhibition even in patients with diabetes and hypertension, we do not anticipate empirical treatment with ACE-I or ARB solely based on presence of coronary atherosclerosis would yield plaque stabilization. Furthermore, we observed higher proportion of patients on high-intensity statins in patients on RAAS inhibitors. This trend is in concordance with recent guidelines advocating for aggressive lipid lowering with high-intensity statin therapy in patients with T2DM and hypertension.^20; 21^ As high-intensity statins have been shown to possess pleotropic effects that can stabilize coronary plaques,^10; 22^ aggressive uptitration of these medications should be emphasized in this high-risk cohort.

The number of patients on tissue specific ACE-I is low (n=4) in our study. Hoshida et al. showed an increased ACE activity in culprit lesions from acute coronary syndrome through analysis of coronary plaques obtained by coronary atherectomy.^23^ Ramipril exerts greater tissue activity than other ACE-I and the HOPE trial demonstrated lower atherosclerotic events with ramipril in patients with vascular disease or diabetes plus one other atherosclerotic risk factor.^17^ Quinapril, another tissue specific ACE-I, has previously shown to improve endothelial dysfunction in normotensive patients.^24^ However, it has not been proven that tissue specific ACE-I is superior to serum ACE-I in lowering major adverse cardiovascular outcomes. Further studies are needed to see whether tissue specific ACE-I would yield greater anti-inflammatory effect on coronary plaques.

In addition to ACE-I and ARB, the other classes of antihypertensive therapies were not associated with lower coronary inflammation on CCTA. Thus far, no study has been done to evaluate how these drug therapies would modulate coronary plaque progression and inflammation on CCTA. Beta blockers historically were the standard of care in medical therapy for coronary artery disease but recent studies showed a lack of long-term clinical benefits in patients who previously experienced acute myocardial infarction and had preserved ejection fraction.^25; 26^ Furthermore, calcium channel blockers, aldosterone receptor antagonists, and thiazides have not shown to yield specific benefits in preventing future myocardial infarction.^27-29^ As such, it is plausible that these treatments are not associated with lower coronary inflammation on CCTA.

Our study has several limitations. It is a single-centered, retrospective observational analysis and falls short of the robust methodology of a randomized, prospective clinical trial. We were unable to verify medication compliance beyond clinical documentations. As some studies demonstrated an association between poor glycemic control and medication adherence,^30; 31^ we excluded patients with HbA1C >9.0%. Furthermore, while we recorded blood pressure readings in the 3 clinic visit encounters preceding the CCTA, we were unable to confirm whether proper blood pressure monitoring techniques were used. While there is good interobserver variability with PCAT attenuation measurement and we eliminated CCTAs with poor imaging quality that could skew the assessment,^32; 33^ it should be noted that the PCAT assessment was done by only one cardiovascular imaging specialist. Our study findings are more applicable for T2DM patients with milder degree of renal involvement, yet the results are still suggestive of greater anti-inflammatory potential associated with ACE-I and ARB in patients with GFR<90 mL/min. Further analyses are warranted to assess the anti-inflammatory potential of these classes of drugs in a population with a broader spectrum of renal failure and alternative imaging modalities that do not require intravenous contrast enhancement, such as positron emission tomography, may serve better for this purpose. Finally, only 16% of our study cohort were found to have severe, obstructive coronary artery stenosis that warrant intervention after CCTA. While we did not observe a trend, further studies are warranted to see if RAAS inhibitors would yield greater anti-inflammatory effect in patients with more advanced coronary artery disease.

## Conclusion

In our cohort of T2DM patients with coronary atherosclerosis, we did not find an independent association between ACE-I, ARB, or any class of first-line antihypertensive therapy and lower coronary inflammation on CCTA. However, those with reduced GFR may receive greater anti-inflammatory benefits from ACE-I or ARB, and large-scale, prospective studies are needed to explore these findings in a wider spectrum of chronic kidney disease.

## Data Availability

Research data contain private patient information and cannot be shared.

## Non-standard Abbreviations and Acronyms

ACE-I: angiotensin-converting enzyme inhibitors
PCAT: pericoronary adipose tissue
CCTA: coronary computed tomography angiography
RAAS: renin-angiotensin-aldosterone system

## Acknowledgments

None

---

**Table 1.** Baseline characteristics of the two treatment groups. Statistically significant differences are highlighted in black. Acronyms-ACE-I/ARB: angiotensin-converting enzyme inhibitors or angiotensin receptor blockers, BMI: body mass index, CAC: coronary artery calcification, HbA1C: hemoglobin A1C, CT: computed tomography, NSTEMI: non-ST elevation myocardial infarction, GLP-1: glucagon-like peptide, SGLT-2: sodium-glucose cotransporter.

| Variables | Not on ACE-I/ARB (n=101) | ACE-I/ARB (n=122) | p-values |
| --- | --- | --- | --- |
| Age | 64.0±8.9 | 65.6±8.7 | 0.164 |
| BMI | 27.7±4.0 | 28.5±4.1 | 0.136 |
| **CAC score** | **518.8±781.3** | **759.5±1026.1** | **0.048** |
| HbA1C (%) | 6.7±0.9 | 6.8±0.8 | 0.519 |
| **Glucose (random, mg/dL)** | **158.2±67.4** | **139.5±3.8** | **0.017** |
| Glomerular filtration rate (mL/min) | 87.9±19.0 | 84.3±17.4 | 0.148 |
| Creatinine (mg/dL) | 0.90±0.28 | 0.91±0.24 | 0.803 |
| Sex (male) | 74.3% | 64.8% | 0.146 |
| Race |  |  |  |
| Caucasians | 35.6% | 32.0% | 0.572 |
| East Asians | 20.8% | 22.1% | 0.871 |
| African Americans | 4.0% | 5.7% | 0.758 |
| Hispanics | 15.8% | 22.1% | 0.306 |
| South Asians | 15.8% | 13.1% | 0.571 |
| Pacific Islanders | 4.0% | 3.3% | 1 |
| Others | 4.0% | 1.6% | 0.414 |
| **Hypertension** | **75.2%** | **97.5%** | **<0.001** |
| **Hyperlipidemia** | **82.2%** | **92.6%** | **0.023** |
| Smokers (current or former) | 32.7% | 33.6% | 1 |
| Microvascular complications | 40.6% | 47.5% | 0.344 |
| Coronary interventions required | 14.9% | 17.2% | 0.716 |
| Indications for CT |  |  |  |
| Chest pain or anginal equivalent | 57.4% | 64.8% | 0.272 |
| **Preoperative evaluation** | **21.8%** | **9.8%** | **0.015** |
| Abnormal cardiac testing | 13.9% | 13.1% | 1 |
| Cardiomyopathy evaluation | 6.9% | 4.1% | 0.385 |
| NSTEMI | 0.0% | 4.1% | 0.065 |
| Other | 0.0% | 4.1% | 0.065 |
| Medications |  |  |  |
| Metformin | 89.1% | 95.1% | 0.128 |
| GLP-1 receptor agonist | 13.9% | 26.2% | 0.204 |
| SGLT-2 inhibitors | 18.8% | 18.9% | 0.368 |
| Insulin | 22.0% | 15.6% | 0.228 |
| Aspirin | 36.6% | 44.3% | 0.275 |
| **High-intensity statins** | **23.8%** | **44.3%** | **0.001** |

